# REPEATABILITY OF A DIGITAL IMAGING TECHNOLOGY FOR DENTAL PLAQUE QUANTITATION (QRAYCAM™ PRO) IN CURRENT, FORMER AND NEVER SMOKERS – STUDY PROTOCOL

**DOI:** 10.1101/2021.01.12.21249560

**Authors:** Gianluca Conte, Sebastiano Antonio Pacino, Toti Urso, Rosalia Emma, Pasquale Caponnetto, Fabio Cibella, Riccardo Polosa

## Abstract

Although the detrimental health effects of smoking on human health are well described, the impact of smoking on dental plaque build-up lacks consistent records. This is because dental research on periodontal health has primarily relied on subjective indices with poor discriminatory power. Novel digital imaging techniques for the objective quantitation of dental plaque are now available. Quantitative Light-Induced Fluorescence (QLF) technology has been used in several studies for digital quantification and monitoring of dental plaque.

The objective of the study is to quantitate and compare short- and long-term repeatability of dental plaque among current, former, and never smokers by using a high resolution, auto-focus, hand-held QLF scanner (QRayCam™ Pro; Inspektor Research Systems BV, Amsterdam, NL).

Demonstration of good reproducibility of QLF technology with clear discrimination for dental plaque quantitation among current, former, and never smokers will pave the way to future application of this test for both medical and regulatory research applied not only to combustion-free tobacco products (e.g. e-cigarettes, heated tobacco products, oral tobacco/nicotine products, etc.) and smoking cessation medications, but also to consume care product for oral hygiene.

## INTRODUCTION

Dental plaque is an oral biofilm (i.e. mass of bacteria) that grows on the tooth surface, typically along the gumline, or below the gumline margins (1). Build-up and maturation of dental plaque give rise to dental calculi, which requires dental scaling for removal (2). Poor oral hygiene (i.e. lack of regular and thorough toothbrushing) is generally associated with dental plaque formation and build-up. The correlation between oral biofilm and periodontal diseases was first established by Loe et al. (3), who were the first to suggest a pathogenetic role for bacteria in dental plaque for the initiation of periodontal inflammation. Therefore, the regular removal of the oral biofilm is one of the most effective ways for preventing periodontal disease (4). Local and systemic risk factors for dental plaque build-up and periodontal disease are well established (5-15). Moreover, periodontitis is an independent risk factor for cardiovascular disease (16) and reducing periodontal disease is likely to have an overall positive impact on cardiovascular health.

The detrimental health effects of cigarette smoking are well known, with several diseases and conditions affecting nearly every organ and system of the human body (17,18). Regular smokers appears to be prone to more severe periodontal manifestations - even after adjusting for age, education level, history of diabetes, BMI, alcohol consumption, perceived mental stress and oral hygiene levels (19). The impact of smoking on dental plaque build-up lacks consistent documentation. For example, in a small short-term study, the rate of plaque formation was similar between smokers and non-smokers (20). The poor discriminatory power of the Silness and Löe plaque index due to the subjective component of investigator scoring, is the most likely explanation for the lack of difference in dental plaque indices between smokers and non-smokers in this study.

Cutting-edge approaches for the quantitation of dental plaque changes are urgently needed to clarify the impact of tobacco smoking on oral health and to improve the quality of study endpoints of oral health in clinical trials. Objective, quantitative dental plaque assessment can be attained by digital imaging techniques after using disclosing solutions or tablets. Carter et al (21), were the first to develop an automated method based on digital imaging of methylene blue disclosed plaque. The method was clearly superior compared to the assessment by dental plaque indices, but lacked careful standardization, reproducibility was not established, and the procedure was quite cumbersome.

More recently, Quantitative Light-Induced Fluorescence (QLF) has been used in several studies, both in vitro (22,23) and in vivo (24,25) and seems to be a promising technique for the digital quantification and monitoring of dental plaque. QLF does not require any additional disclosing procedure as it measures dental plaque by quantitively analyzing the red fluorescence from dental plaque induced by blue visible light at a wavelength of 405 nm. Reproducibility of QLF is required to be confident of test results and no data is available about its short- and long-term reproducibility.

The issue of test variability is particularly important when investigating subjects with poor dental hygiene or with gum disease or with significant exposure to tobacco smoke. No data is available about changes of QLF in smokers who quit smoking. The impairment of dental plaque caused by smoking may be permanent with little possible restoration after smoking abstinence. If this is true, former smokers should exhibit similar QLF values as in current smokers.

Objective of the study is to assess and validate a novel QLF technology by quantitating and comparing short- (7 days) and long-term (30 days) repeatability of dental plaque among current, former, and never smokers by using digital imaging technology consisting of a high resolution, auto-focus, hand-held QLF scanner (QRayCam™ Pro; Inspektor Research Systems BV, Amsterdam, NL).

## METHODS

### Study Population

The study population will consist of three study groups identified among a pool of subjects who attended a smoking cessation clinic (CPCT, Centro per la Prevenzione e Cura del Tabagismo of the University of Catania) in the previous 2 yrs or contacted among hospital staff.

Study group 1 consisted of current smokers, defined as smokers of > 10 cigarettes per day with an exhaled carbon monoxide (eCO) level of ≥7 ppm.

Study group 2 was formed of former smokers, defined as quitters of at least 12 months and who were still abstinent when contacted for enrollment, with an eCO level of < 7 ppm.

Study group 3 consisted of never smokers, defined as having never smoked or who reported having smoked less than 100 cigarettes in their lifetime (26). Their eCO had to be < 7 ppm to exclude subjects significantly exposed to cigarette smoke or to environmental sources of carbon monoxide.

Current, former and never smokers had to satisfy the following **inclusion criteria**:

- healthy adult subjects (age 18-50 yrs), with no systemic and/or chronic disease
- presence of at least 10 natural anterior teeth (cuspid to cuspid, lower and upper jaw), with no prosthetics or crown

Furthermore, they had to satisfy the following **exclusion criteria**:

∘ Any conditions that could interfere with dental plaque measurements, including:
  ▪ Recent (less than 30 days) hx of oral/dental infection/inflammation
  ▪ Recent (less than 15 days) course of antibiotics or antinflammatory drugs
  ▪ Subjects wearing fixed or removable orthodontic appliance or prothesis (limited to the 12 natural anterior teeth);
  ▪ Significant exposure to passive smoking (excludes current smokers)
∘ Had undergone dental professional cleaning (i.e. scale and polish) within 6 months prior to screening
∘ Pregnancy

### Study Design

This is an observational study with a cross-sectional design to assess dental plaque by QLF imaging among three study population (current, former, and never smokers). The study consists of a total of four visit: screening visit, baseline visit at day 0 (Visit 1), short-term follow-up visit at day 7 (±1 days) (Visit 2) and long-term follow-up visit at day 30 (±3 days) (Visit 3) (Figure 1). Participants will be asked:

**Figure 1.**
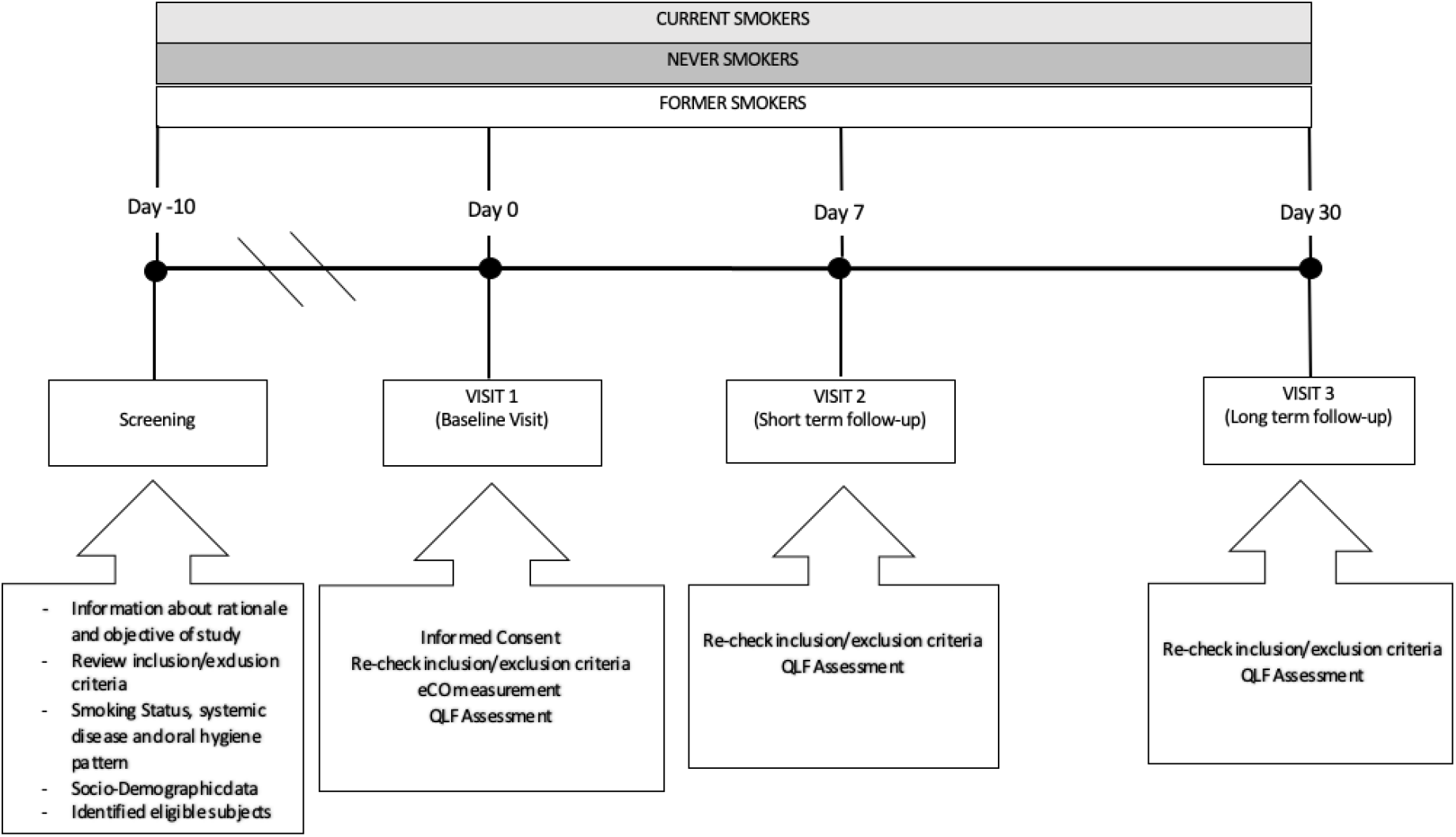
Schematic diagram of the study design

- not to change their own habitual oral hygiene (mouthwash, mouth rinse, interdental-floss, ecc.) pattern for the whole duration of the study
- avoid scaling and polishing procedures for the whole duration of the study not to floss for at least 72 hrs prior to each study visit
- not to smoke for at least 2 hrs prior to each study visit
- not to tooth brush and/or mouth rinse for at least 2 hrs prior to each study visit
- not to eat and/or drink (except water) for at least 2 hrs prior to each study visit

### Study Visits

#### Screening Visit

Potential participants will attend a screening visit to 1) receive information about the rationale and objectives of the research; 2) verify eligibility criteria by reviewing their inclusion and exclusion criteria; 3) assess smoking status and oral hygiene habit (i.e. frequency of toothbrushing, type of toothpaste, etc.); and 4) record general socio-demographic characteristics (i.e. sex, age and occupation). All eligible subjects will be invited to participate to Baseline Visit (Visit 1).

#### Baseline Visit (Visit 1)

The Baseline Visit will be carried out within 10 days of the Screening Visit. Participants will be asked to go over a patient information sheet and to sign a consent form. After re-checking inclusion/exclusion criteria and reviewing study restrictions, eCO measurement and QLF assessment will be carried out and baseline data recorded. Participants will be instructed not to change their habitual oral hygiene pattern and invited to attend next study visit (Visit 2).

#### Day-7 Visit (Visit 2)

Visit 2 will be carried out within 7 (±1) days after Visit 1. Eligibility criteria will be verified again. QLF assessment will be repeated for short term repeatability. Participants will be instructed not to change their habitual oral hygiene pattern and invited to attend next study visit (Visit 3).

#### Day-30 Visit (Visit3)

Visit 3 will be carried out within 30 (±3) days after Visit 1. After re-checking eligibility criteria, QLF assessment will be repeated for long term repeatability.

#### Exhaled Carbon Monoxide Measurement

The smoking status will be objectively verified by measuring exhaled carbon monoxide (eCO) levels (eCO > 7 ppm indicating smoking status) with a portable CO monitor (Micro CO; Micro Medical Ltd, UK). Participants will be asked not to smoke any cigarettes for at least 2 hours prior to eCO level measurements. Participants will be invited to exhale slowly into a disposable mouthpiece attached to the eCO monitor as per manufacturer’s recommendations. The value of eCO readings will be noted.

#### QLF Assessment

Prior to QLF assessment, participants will be asked to rinse their mouth with water and subjected to gentle flushing and drying by triple syringe tip to remove any food debris. Saliva will be minimized both by asking participants to swallow and eventually using a dental vacuum. Cheek retractors (Henry Schein, Gillingham, UK, Double end large, 106-7079) will be placed to allow a good view of the 12 natural anterior teeth.

The Q-ray cam Pro (Co., AIOBIO, Seoul, Republic of Korea) is a high resolution, lightweight, handheld and auto-focus QLF™ camera (Figure 2). Images of the anterior teeth will be taken with the Q-ray cam™ Pro according to the standard QLF digital photography protocol. The camera will be brought as close to the subject’s teeth as physically possible. Ambient light level should be standardized, making sure that any excessive ambient light is avoided. The auto-focus function of the camera will be used to focus on the maxillary lateral incisor and canine (focal depth 0.32) and a white-light and a QLF image will be taken automatically in quick succession and will be recorded by the Q-Ray™ software (QA v.1.41, Inspektor Research Systems BV, Amsterdam, NL). Q-ray cam Pro will be used with the following settings: Resolution (image size), full high-definition [1920*1080 pixels]; shutter speed, auto [1/30–1/30000 s]; aperture, Auto [F1.2–360]; sensor object distance, 2.3Mpixel Image Sensor.

**Figure 2.**
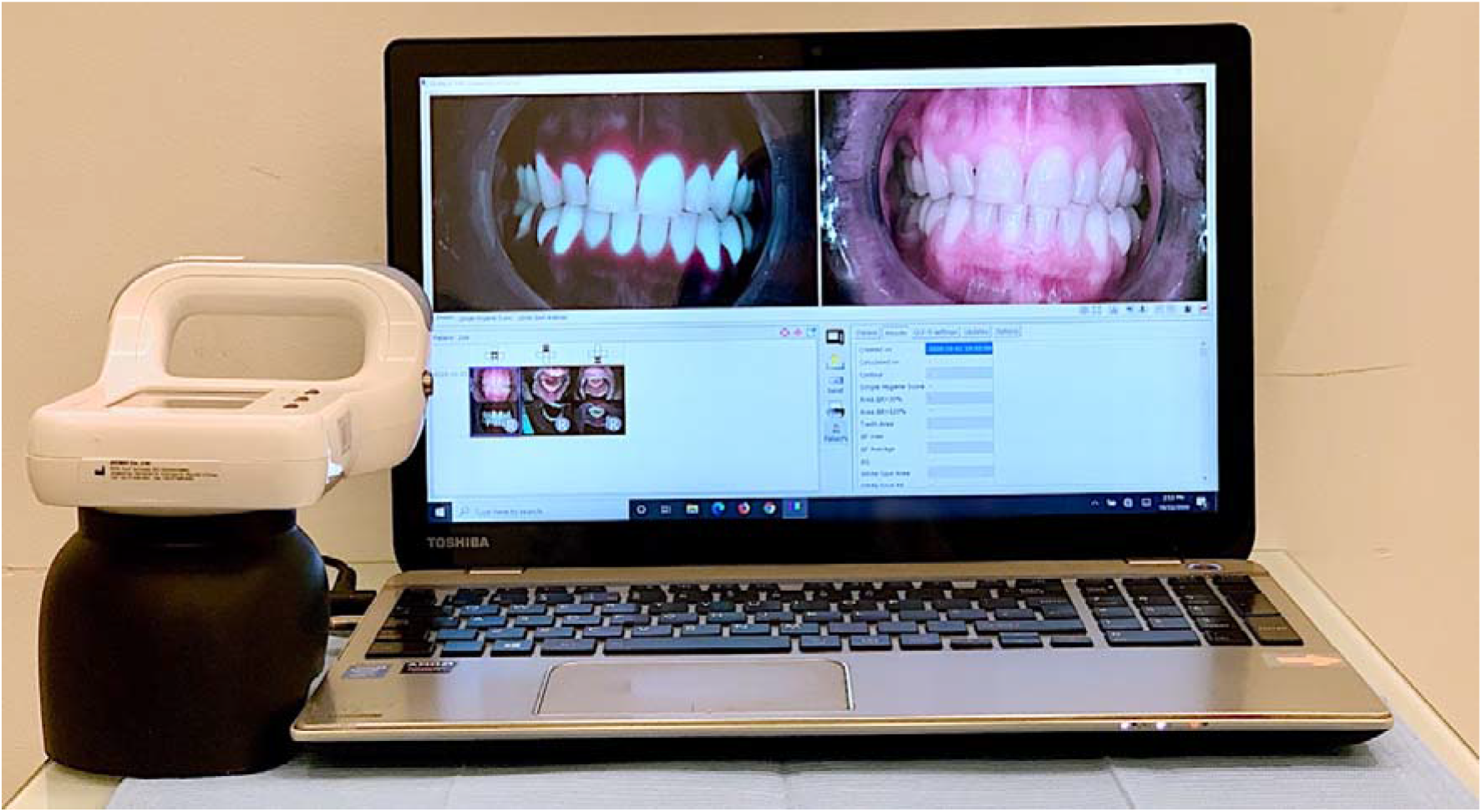
Q-ray cam Pro connected to a portable computer with pre-installed Q-Ray™ software.

Fluorescence photographs of the vestibular aspect of the anterior teeth (cuspid to cuspid, upper and lower jaw) in end-to-end position will be taken. Images will be automatically saved by default as a bitmap image using a QLF proprietary software (QA v.1.41, Inspektor Research Systems BV, Amsterdam, NL). The software will analyze the images to determine, for each pixel on the dental surface, the value of ΔR which is a measure of the increase of red fluorescence relative to the sound surface. Higher ΔR values indicate areas with more active bacterial metabolism within the dental plaque, representing a greater level of dental plaque maturation. Therefore, the percentage of the total tooth surface showing an increase in the intensity of at least 30% (ΔR30) indicates the total area of mature dental plaque detected, whereas an increase of 120% (ΔR120) reveals only areas with greater level of plaque maturation (i.e. calculus/tartar) within the total area of mature dental plaque detected.

#### Data Analysis

This will be a proof-of-concept pilot study, the first of its kind, hence no previous data could be used for power calculation.

Short-term repeatability of the QLF assessment will be evaluated by linear regression analysis of measurements obtained at V1 and those obtained at V2 for each study group. Likewise, long-term repeatability will be evaluated by linear regression analysis of measurements obtained at V1 and those obtained at V3. Scatter plots of linear regression analyses will be generated to visualize repeatability results. Moreover, “Bland and Altman” plots will be created to describe the level of agreement between V1 vs V2 and V1 vs V3 for each study groups. 1-tailed sample t test will be also performed to assess the difference from zero of the mean difference between two measurements.

The Range of Normality will be calculated by computing the value corresponding to the mean ± SD*1.64 from the distribution curve of the results of the QLF measurements in never smokers. Kolmogorov-Smirnov test will be performed to assess the normality data. Categorical data will be summarized by counts and percentages; continuously distributed data, with symmetrical distribution, will be summarized using the mean (standard deviation; SD); continuously distributed data, with skewed distribution, will be summarized using the median (inter-quartile range; IQR). Study groups comparisons will be carried out by Chi-square test, ANOVA and Kruskall-Wallis test for categorical, continuously symmetric and continuously skewed datasets, respectively.

Multiple regression tests will be also performed to identify individual variables, including age, gender, eCO level, pack/years, cig/day, type of toothbrush (i.e. electric toothbrush), type of toothpaste, frequency of toothbrushing, etc. that may influence the results of the QLF assessment. All analyses will be considered significant with a P-value< 0.05. R version 3.4.3 (2017-11-30) will be utilized for data analysis and generation of graphs.

## RESULTS

Recruitment of participants started in July 2020 and was completed in November 2020. Final results will be reported in 2021.

## DISCUSSION

The evidence about the impact of regular smoking on dental plaque build-up and gingival bleeding is inconsistent, because dental research has primarily relied on highly variable and poorly sensitive subjective scoring indices with poor discriminatory power.

Several dental plaque indices have been developed to try and improve the limitations of quantitating dental plaque by earlier scoring indices, but failing to achieve consistent findings in spite of similar study designs (27-33).

The results of these studies are limited by the subjective nature of the dental plaque indices, considering that they are all based on visual examination. Visual indices can be improved when examiners are properly training, but inter-examiner consistency is hard to achieve.

Innovative 21st century technologies are now needed for objective and consistent assessment of the dental plaque. Novel planimetric digital imaging and analysis techniques for the objective quantitation and monitoring of dental plaque are now available including Quantitative Light-Induced Fluorescence (QLF) technology, which is non-invasive, well tolerated, and simply to perform.

In this pilot study we intend to verify whether QLF technology (QRayCam™ Pro; Inspektor Research Systems BV, Amsterdam, NL) can consistently measure and detect dental plaque. Standardization and reproducibility of the QLF technology are required to minimize its variability and to be confident of test results.

The issue of test variability is particularly important when investigating subjects with poor dental hygiene or with gum disease or with significant exposure to tobacco smoke. Moreover, no information is available about changes of QLF in smokers who quit smoking. Acceleration in dental plaque build-up caused by smoking may be permanent with little possible reversal after smoking abstinence. If this is true, former smokers should exhibit similar QLF results as in current smokers.

Hopefully, demonstration of good reproducibility of QLF technology with clear discrimination for dental plaque quantitation among current, former, and never smokers will have an important impact on future application of this test for both medical and regulatory research applied not only to combustion-free tobacco products (e.g. e-cigarettes, heated tobacco products, oral tobacco/nicotine products, etc.) and smoking cessation medications, but also to consume care product for oral hygiene.

